# Deep Learning Chest X-Ray Age, Epigenetic Aging Clocks and Associations with Age-Related Subclinical Disease in the Project Baseline Health Study

**DOI:** 10.1101/2025.01.02.24319734

**Authors:** Jay Chandra, Sarah Short, Fatima Rodriguez, David J. Maron, Neha Pagidipati, Adrian F. Hernandez, Kenneth W. Mahaffey, Svati H. Shah, Douglas P. Kiel, Michael T. Lu, Vineet K. Raghu, the Project Baseline Health Study Group

## Abstract

Chronological age is a cornerstone of medical decision-making but is limited because individuals age at different rates. We recently released an open-source deep learning model to assess biological age from chest radiograph images (CXR-Age), which predicts incident all-cause and cardiovascular mortality better than chronological age. Here, we compare CXR-Age to two established epigenetic aging clocks (First generation – Horvath Age; Second generation - DNAm PhenoAge), to test which is more strongly associated with measures of cardiopulmonary disease. Our cohort consisted of 2,097 participants from the Project Baseline Health Study (PBHS), a prospective cohort study of individuals from four US sites enriched for cardiovascular and cardiometabolic disease risk factors. We found that CXR-Age was most strongly associated with the presence of coronary calcium, cardiovascular risk factors, worsening pulmonary function, increased frailty, and abundance in plasma of two proteins implicated in neuroinflammation and aging. Associations with second generation epigenetic clocks were weaker for pulmonary function and for all outcomes in younger adults. No associations were found with first generation clocks. These results suggest that opportunistic screening using CXR-Age may help identify high risk individuals who could benefit from directed screening and prevention.

## INTRODUCTION

Chronological age is a critical component of medical risk scores (e.g., atherosclerotic cardiovascular disease risk calculator)^1^ and medical decision-making (e.g., 50-77 years for lung cancer screening);^2^ however, there is considerable variability in how individuals age.^3^ Accurate measures of “biological age” can improve medical decisions currently predicated on chronological age. Tools to measure biological age (“biological aging clocks”) aim to summarize accumulated molecular and cellular damage and commensurate functional decline into a single number that matches chronological age in the average individual.^4^ Aging clocks have been derived from molecular sources (epigenetics,^5^ proteomics,^6^ telomere length);^7^ functional performance (grip strength,^8^ gait speed);^9^ and physical characteristics (waist circumference^10^, and muscle and fat composition).^11^

Of these, the most studied clocks are epigenetic clocks which predict chronologic age based on methylation levels at CpG sites across the genome. First-generation epigenetic clocks were developed to strongly correlate to chronological age (r∼0.9) using methylation patterns across a variety of tissues^12^ and whole blood.^13^ The deviation between blood-based epigenetic clocks and chronological age (termed “age acceleration”) is associated with cancer, dementia, and all-cause mortality.^14^ More recent “second-generation” clocks (e.g., DNAm PhenoAge) have stronger associations with disease by focusing on methylation sites associated with abnormal clinical biomarkers.^15^ This DNAm PhenoAge clock was more strongly associated with mortality due to age-related disease (e.g., heart disease, cancer, lower respiratory disease, etc.) and age-related biomarkers (cholesterol, glucose, systolic blood pressure, etc.) than first-generation clocks targeted to predict chronological age.^15^

Biological aging is complex, and multiple aging clocks from different data sources may accurately measure components of this process.^4^ Methylation clocks themselves can give discordant predictions^16^, and multiple clocks with similar accuracy can be found using distinct CpG sites,^17^ suggesting that each captures only a component of the aging process. For clinical use, another main limitation of methylation clocks is that they require epigenetic testing not routinely acquired in practice.^18^

Radiographic clocks from routine medical imaging are a promising way to measure a component of biological aging using images acquired in routine clinical practice. In particular, chest radiographs (CXRs) are among the most common tests in medicine and existing CXRs may enable biological age measurement without additional testing.^19^ We recently released a convolutional neural network (CNN) model to assess biological age from chest radiograph images (CXR-Age) (https://github.com/circ-ml/CXR-Age).^20^ The CXR-Age model was first trained to predict chronological age using ∼25,000 healthy patients. It was then fine-tuned on ∼15,000 individuals’ images from the Prostate, Lung, Colorectal, Ovarian (PLCO) Cancer Screening Trial to predict a phenotypic age based on chronological age, prevalent risk factors, and observed mortality over 18-years of follow-up. This model was validated using ∼40,000 other PLCO participants, ∼5,000 National Lung Screening Trial participants, and 36,924 individuals undergoing health checkups in South Korea.^21^ In all cohorts, CXR-Age was associated with all-cause, cardiovascular, and respiratory mortality beyond chronological age and prevalent risk factors.

Limitations of these studies were the absence of information on subclinical disease measures, no direct comparison of CXR-Age with known measures of biological aging, and lack of interpretability in how the CXR-Age model produces a biological age output. Here, we compare CXR-Age with first (Horvath Age) and second-generation (DNAm PhenoAge) epigenetic clocks by assessing their association with age-related disease processes including cardiovascular disease, lung function, frailty, and disability. We then conduct plasma proteomic analyses to better understand potential mechanisms captured by the CXR-Age model and epigenetic clocks.

## RESULTS

### Cohort Characteristics

The study cohort consisted of 2,097 people who had CXRs as part of the Project Baseline Health Study^22^ (mean age 51.5 years, 56.2% female; Table 1). 61.6% of participants were Caucasian, 17.6% Black or African American, and 20.8% were another self-reported race. Among the 1,294 who also had DNA Methylation data, the mean CXR-Age in the cohort was 53.1 (± 3.5 sd), mean Horvath age was 55.2 (± 14.2), and mean DNAm PhenoAge was 39.1 (± 15.9). The mean CXR-Age acceleration was 0.0 (±3.5), mean Horvath age acceleration was -0.05 (±4.3), and mean DNAm PhenoAge acceleration was -0.2 (±5.8) (Supplemental Figure 1).

**Table 1.** Project Baseline Health Study (PBHS) cohort characteristics.

|  |  | **Project**<br>**Baseline** |
| --- | --- | --- |
| N |  | 2097 |
| CXR-Age (years) |  | 53.1 (3.5) |
| Horvath DNA methylation Age (years) |  | 55.2 (14.2) |
| DNAm PhenoAge (years) |  | 39.1(15.9) |
| Chronological Age (years) |  | 51.5 (17.0) |
| Log CAC Score |  | 1.7 (2.4) |
| Log ASCVD Risk Score |  | -3.8 (2.0) |
| Sex (%) | Female | 1178 (56.2) |
|  | Male | 919 (43.8) |
| BMI (kg/m2) |  | 28.0 (6.9) |
| Site (%) | Durham, NC | 404 (19.3) |
|  | Kannapolis, NC | 343 (16.4) |
|  | Los Angeles, CA | 457 (21.8) |
|  | Palo Alto, CA | 893 (42.6) |
| Race (%) | American Indian or Alaska Native | 28 (1.3) |
|  | Asian | 199 (9.5) |
|  | Black or African American | 370 (17.6) |
|  | Native Hawaiian or Other Pacific Islander | 22 (1.0) |
|  | Other | 186 (8.9) |
|  | White | 1292 (61.6) |
| Smoking Status (%) | Current | 321 (15.2) |
|  | Former | 455 (21.6) |
|  | Never | 1321 (63.0) |

Values are mean (sd) or percentages.
DNAm – DNA Methylation
CAC – Coronary Artery Calcium
ASCVD – Atherosclerotic cardiovascular disease

### Correlation between CXR-Age, DNA Methylation clocks, and chronological age

There was moderate correlation between CXR-Age and chronological age (r=0.50, 95% CI: [0.47-0.53]), and a high correlation between epigenetic and chronological age (DNAm PhenoAge: r = 0.93 [0.92-0.94], Horvath r = 0.95 [0.94-0.96]) (Figure 1). There was no significant association between CXR-Age acceleration and both DNAm PhenoAge acceleration (β = 0.08 [-0.04 – 0.18]) and Horvath age acceleration (β = -0.02 [-0.11 – 0.06]) after adjusting for covariates including chronological age.

**Figure 1.**
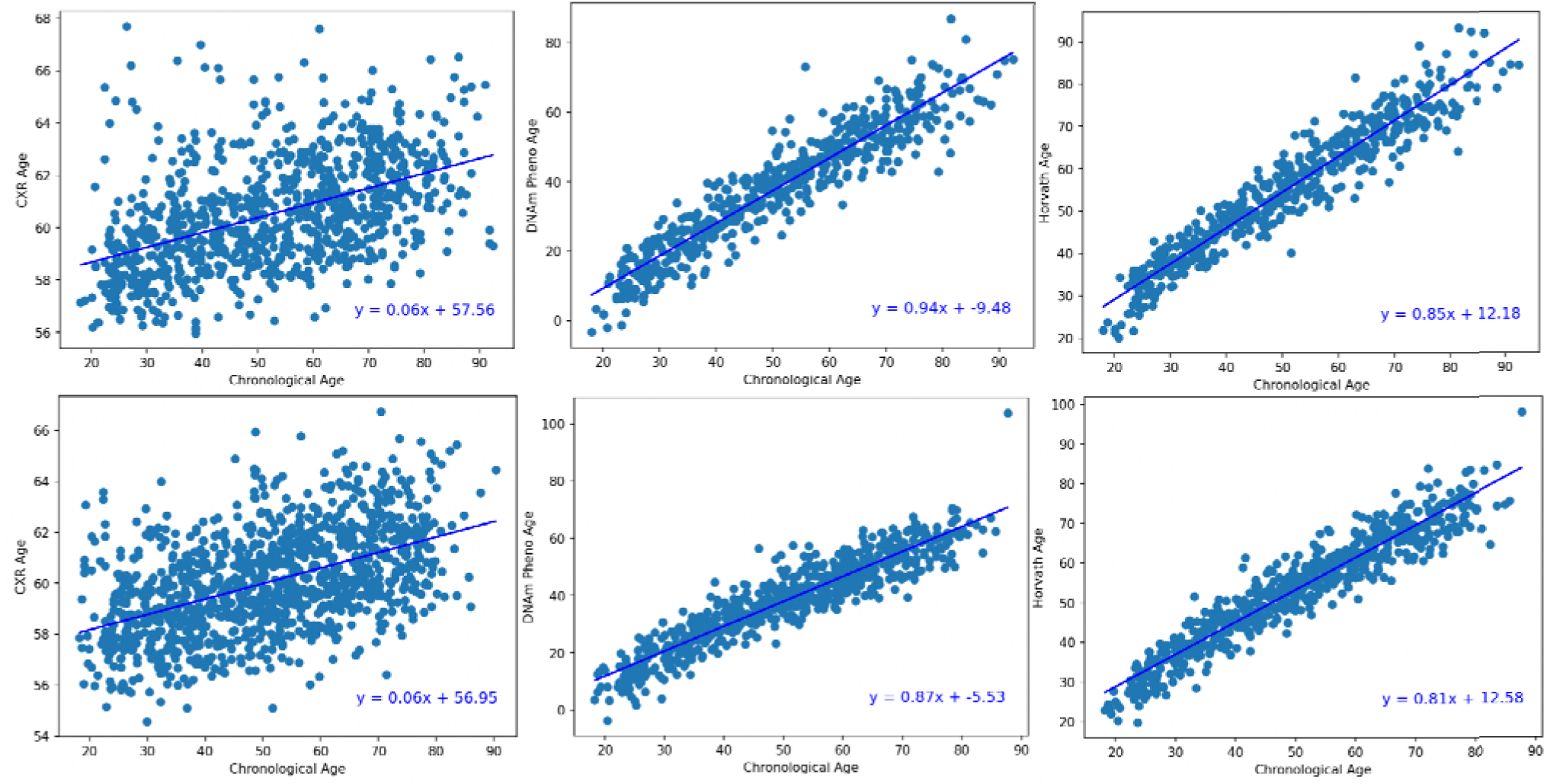
Association of Chronological and Biological Age (CXR-Age, DNAm PhenoAge, Horvath Age) Stratified by male (top) and female (bottom) sex.

### Association of CXR-Age and Epigenetic Clocks with cardiovascular disease

Each year of CXR-Age acceleration was associated with a 1.10-fold [1.06-1.14] increase in coronary artery calcium score (CAC) and each year of DNAm PhenoAge acceleration was associated with a 1.03-fold [1.01-1.05] increase while there was no significant relationship between CAC and Horvath age. When stratifying by chronological age categories (<45,45-60, >60 years), there was a significant relationship between CXR-Age and CAC in the older two groups (Supplemental Figure 2). In all age subgroups, each year of CXR-Age and DNAm PhenoAge was associated with a 1.04-fold [1.01-1.07] and 1.00-fold [0.99-1.01] increase in 10-year ASCVD risk calculated using the Pooled Cohort Equations^23^ based on cardiovascular risk factors. There was not a significant association between Horvath age and ASCVD risk.

### Association of CXR-Age and Epigenetic Clocks with pulmonary function

There was a significant negative relationship between most measures of pulmonary function (FEV1/FVC ratio, DLCO % predicted, FEV1 % predicted-except peak expiratory flow) and CXR-Age acceleration. The strongest association was seen with DLCO % predicted where every 1 year of CXR-Age acceleration was associated with a 0.74% [0.42-1.06] lower DLCO % predicted. For the epigenetic clocks, there was only a significant, negative association between the DNAm PhenoAge score and DLCO% predicted. (Figure 2). When stratifying by chronological age (<45,45-60, >60), there was a significant association between CXR-Age and DLCO% predicted and FVC% predicted in the older two groups (Supplemental Figure 3). When stratifying by ever-vs. never-smokers, we found a significant association between CXR-Age and DLCO% predicted in both groups. For FVC% predicted, there was a significant relationship among never-smokers, and for FEV1/FVC ratio, there was a significant relationship among ever-smokers (Supplemental Figure 4). There was only a significant relationship between DNAm PhenoAge and FEV1/FVC ratio in ever-smokers.

**Figure 2.**
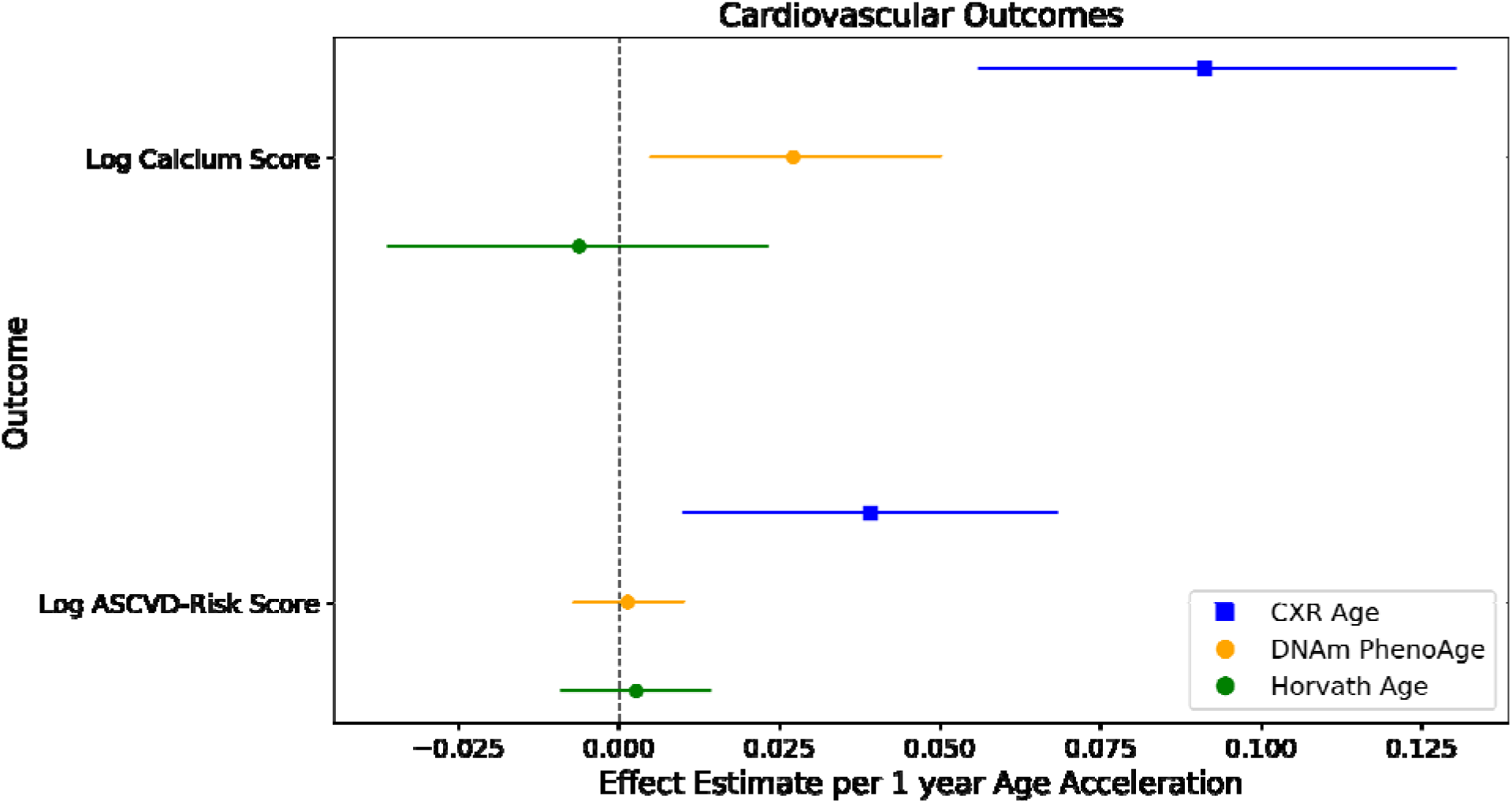
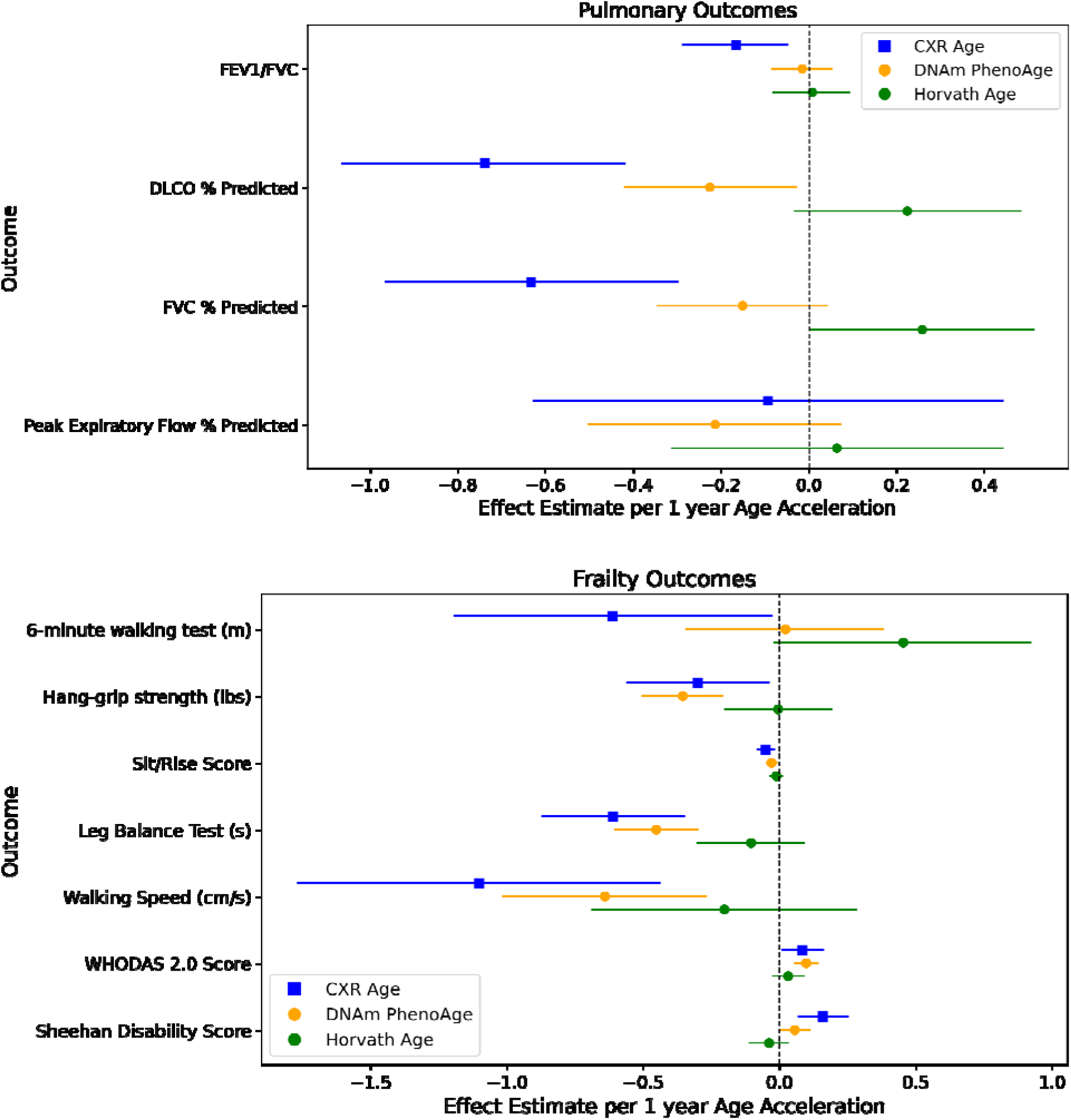
Association of Cardiovascular Outcomes (top), Pulmonary Function Measures (middle), and Frailty Outcomes (bottom) with CXR-Age and Epigenetic Aging Clocks. Effect estimates and 95% CI per 1-year for each age acceleration metric are provided on the x-axis.

### Association of CXR-Age and Epigenetic Clocks with frailty and functional ability

In all five available physical function tests, higher CXR-Age acceleration was associated with lower functional ability. Each year of CXR-Age acceleration was associated with 0.30 [0.04 – 0.56] lbs lower hand grip strength, 0.61 [0.03 – 1.19] meter less six-minute walking distance, 1.1 [0.44 – 1.77] centimeter/second slower walking speed, 0.05 [0.02 - 0.08] lower sit/rise score and 0.61 [0.35 – 0.87] second lower time balancing on a leg (averaged between two legs). Similar, significant associations were found between DNAm PhenoAge acceleration and metrics of frailty and disability. Each year of DNAm PhenoAge acceleration was associated with 0.36 [0.21-0.50] lbs less hand grip strength, 0.45 [0.30 -0.60] fewer seconds balancing on a leg, and 0.64 [0.27 – 1.01] meter/second slower walking speed (Figure 2). CXR-Age and DNAm PhenoAge acceleration were also positively associated with two measures of disability (Sheehan Score and WHODAS 2.0 score; Figure 2). There were no significant associations between Horvath age and the frailty/disability metrics. When stratifying by chronological age (<45,45-60, >60), there was a significant relationship between CXR-Age and the leg balance test, sit/rise score, and the Sheehan disability score in the older two groups (Supplemental Figure 4).

### Plasma proteins associated with CXR-Age and Epigenetic Clocks

Two proteins had a significant, negative relationship with CXR-Age acceleration: CDH13 (T-Cadherin) (0.008 [0.004 - 0.011] decrease in protein abundance per 1-year age acceleration) and ApoD (Apolipoprotein D) (-0.005 [-0.008 - -0.003]). While not significant, SAA2 (Serum Amyloid A2) (0.024 [0.011 – 0.037]) and DBH (Dopamine beta-hydroxylase) (-0.015 [-0.027 – -0.004]) had large, adjusted effect sizes. Thirteen proteins were significantly associated with DNAm PhenoAge acceleration. GSN (Gelsollin) had the lowest p-value and S100A8 had the largest adjusted effect size. CDH13 was significantly associated with both CXR-Age acceleration and DNAm PhenoAge acceleration. There were no significant associations between protein abundance and Horvath age acceleration.

### Interpretation of chest radiographs with high and low CXR-Age acceleration

We manually examined 100 CXRs with the lowest CXR-Age acceleration and 100 CXRs with the highest CXR-Age acceleration. The CXRs with the lowest age acceleration tended to have clearer lung fields and overall were read as “normal” by the interpreting radiologist (Figure 4). In contrast, the CXRs with the highest age acceleration had various lung findings including interstitial infiltrates, patchy infiltrates, pleural effusions, hilar prominence, peribronchial cuffing, evidence of prior thoracic surgery, and pacemaker placement. In some cases, there were no clear findings on the CXR that could correlate to a higher age acceleration.

**Figure 3.**
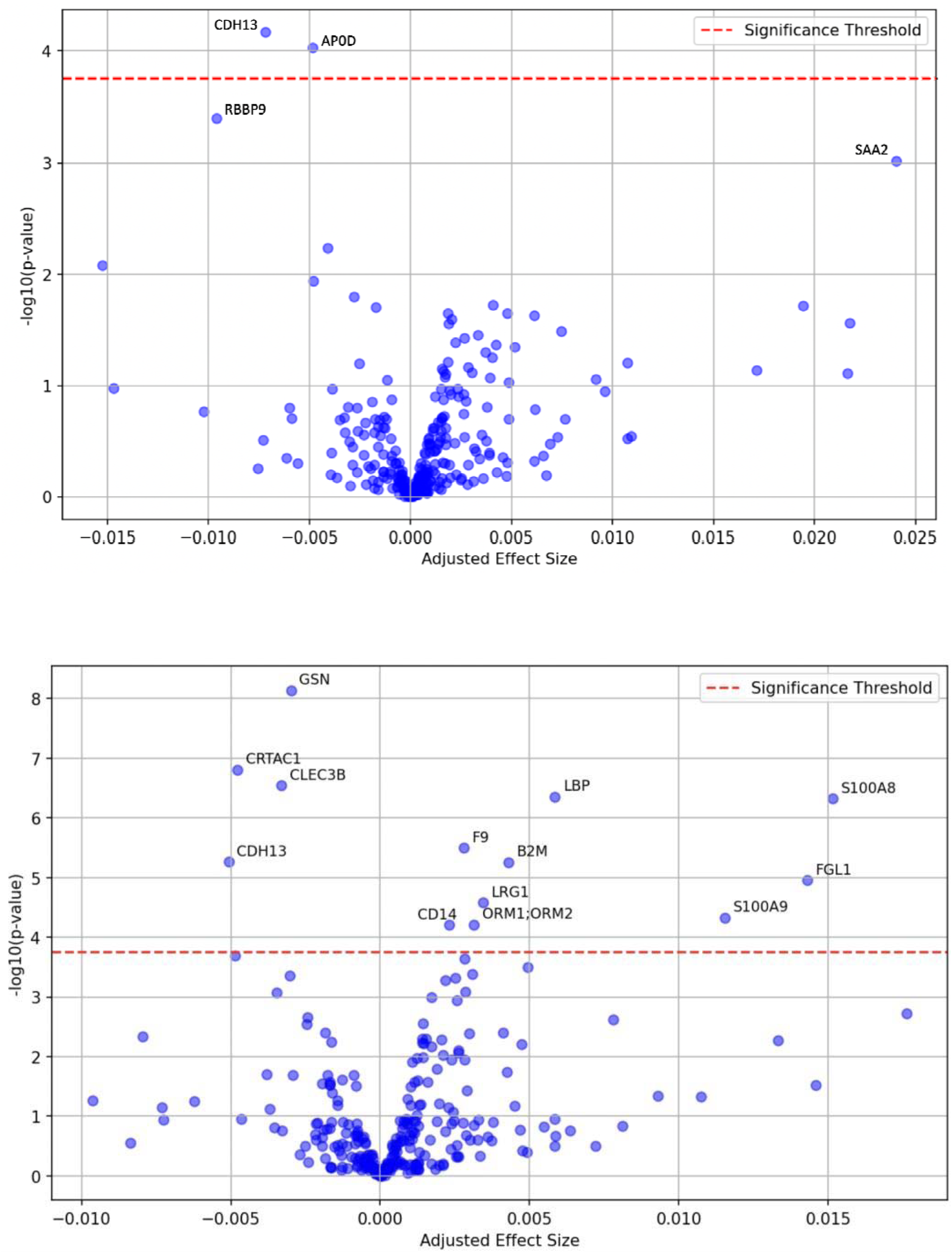
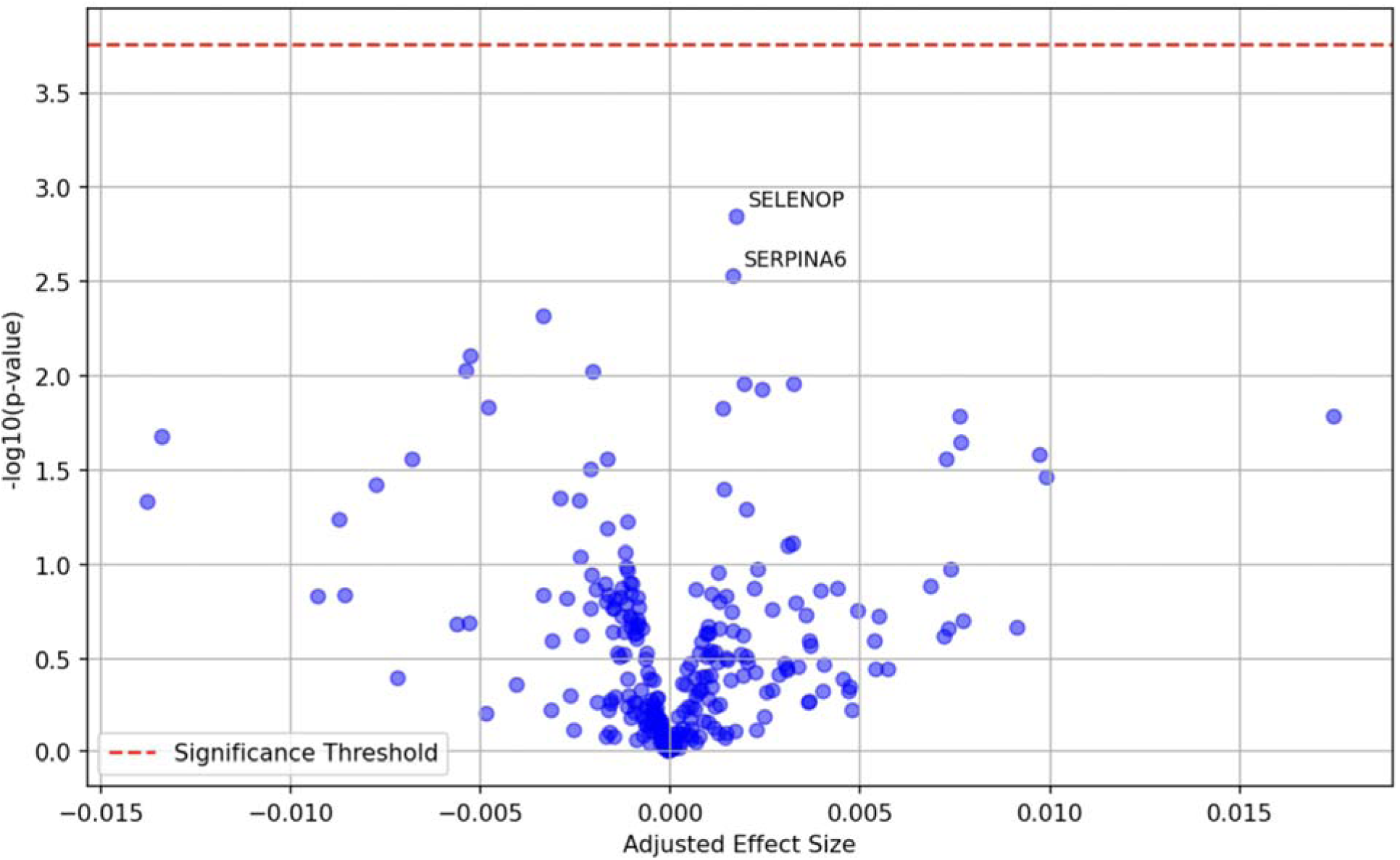
Association between CXR-Age (top), DNAm PhenoAge (middle), Horvath Age (bottom) Acceleration and abundance of 289 proteins in plasma.

**Figure 4.**
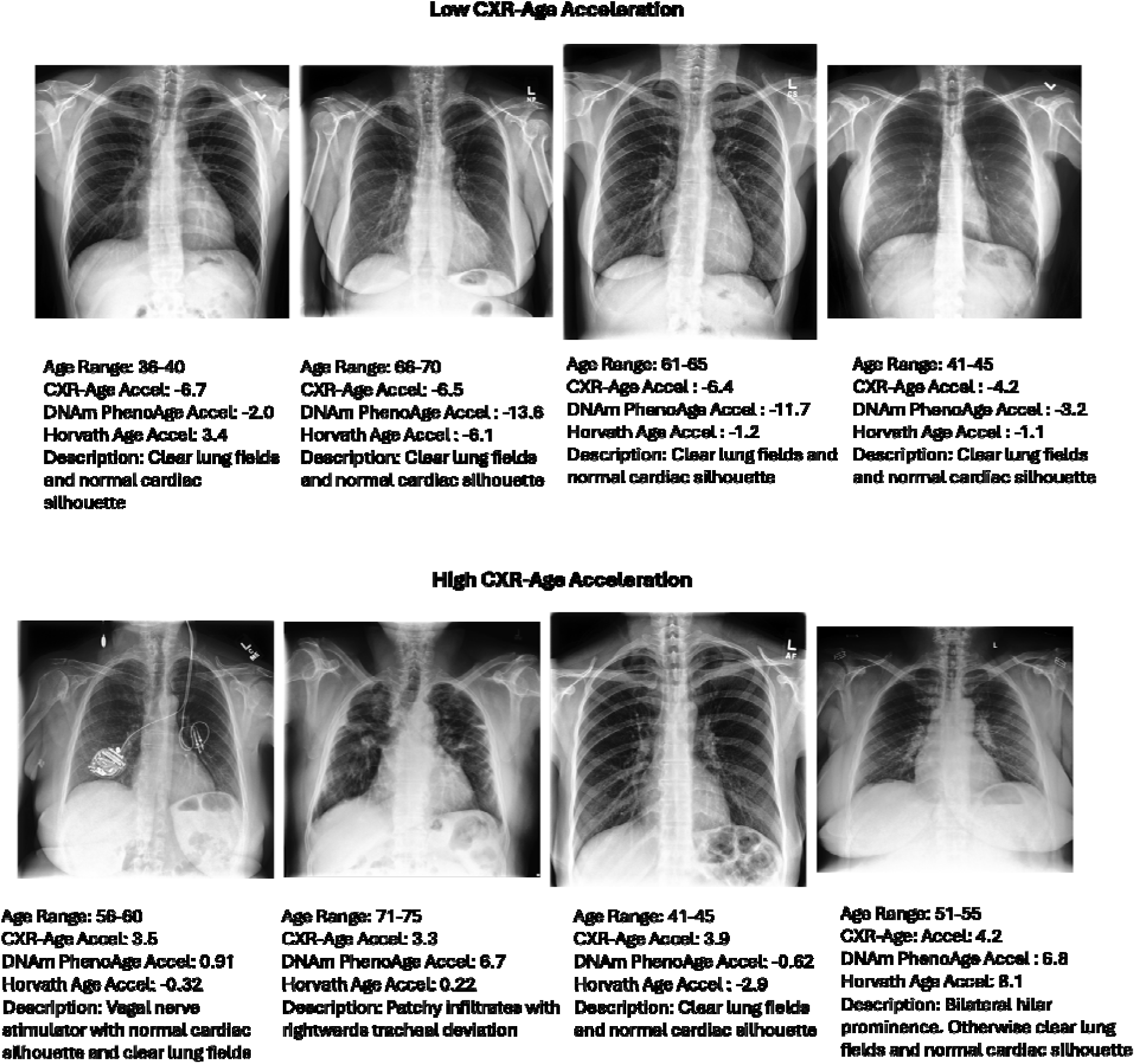
Representative Chest-X-rays with the Lowest CXR-Age Acceleration. CXRs with low age acceleration tended to have clear lung fields and normal cardiac silhouettes, while CXRs with high age acceleration tended to have lung field and airway findings such as patchy infiltrates, hilar enlargement, peribronchial cuffing, and deviated trachea. In addition, in general, the cardiac silhouette seemed to be larger in high age acceleration patients and there was a greater prevalence of hardware including pacemakers.

## DISCUSSION

Chronological age is a cornerstone of medical decision making, but it does not account for individual variability in the rate of aging. Biological aging clocks have the potential to improve medical decisions, but different clocks may capture complementary aspects of biological aging. In this work, we leveraged a diverse sample from the Project Baseline Health Study (PBHS) to assess the association of cardiovascular health, pulmonary function, frailty, and disability with a chest x-ray-based radiologic clock (CXR-Age) and two established epigenetic clocks (Horvath Age and DNAm PhenoAge). Our major findings were 1) chronological age was more strongly associated with epigenetic aging clocks (r ≈ 0.94) than CXR-Age (r=0.49), 2) deviations of the CXR-Age and DNAm PhenoAge clocks from chronological age were associated with subclinical coronary atherosclerosis, abnormal pulmonary function testing, and measures of frailty/disability, 3) the association of CXR-Age with functional measures was robust in adults as young as 45, which was not the case for DNAm PhenoAge, and 4) plasma proteomic analysis revealed novel associations of CXR-Age Acceleration with abundance of two proteins (CDH13 and ApoD) and DNAm PhenoAge with thirteen plasma proteins including CDH13.

CXR-Age is a deep learning-based model that estimates a biological age based on a single chest x-ray image. We previously showed that CXR-Age was moderately associated with chronological age but predicts all-cause and cardiovascular mortality over 18-years of follow-up beyond chronological age and prevalent risk factors in two US cancer screening populations^20^ and in a South Korean health checkup population.^21^. Here, we used the existing CXR-Age model without modification to better understand how CXR-Age estimates biological age. We provide evidence that those at high CXR-Age relative to their chronological age have higher subclinical cardiovascular disease, lower pulmonary function, and higher rates of frailty and disability after adjusting for demographics, smoking, and BMI, suggesting that CXR-Age may be capturing broad aging processes rather than specific pathology.

We found that second generation (time to death predictors; e.g., DNAm PhenoAge) but not first generation (chronological age predictors; e.g., Horvath Age) epigenetic clocks are associated with subclinical cardiovascular disease including CAC; corroborating prior studies. ^24,25^ We further validate prior studies that second generation clocks are associated with disability^26^ but are not strongly associated with a broad set of functional measures^27^. Despite strong associations of second generation clocks with incidence of chronic airflow obstruction^28,29^, prior studies found little association between second generation clocks and pulmonary function tests^29^. In contrast, we found a weak association between DNAm PhenoAge and lung diffusion capacity; however, this was primarily in older adults and ever-smokers and may be confounded by smoking intensity and duration^30^.

Despite concordant associations with cardiovascular disease and disability, there was no significant association between CXR-Age and DNAm PhenoAge, suggesting that they capture different aspects of aging. To gain further insight into aging processes captured by CXR-Age and DNAm PhenoAge, we conducted plasma proteomic analyses and found associations of four plasma proteins with CXR-Age (CDH13, ApoD, SAA2, and DBH) and 13 proteins associated with DNAm PhenoAge, including CDH13, which has known associations with deficits in learning and memory in addition to neurodevelopmental and psychiatric disorders^31^. ApoD is neuroprotective due to its presence in response to oxidative stress and inflammation and is linked to longevity, albeit through an unknown mechanism^32^. SAA2 is strongly associated with chronic inflammatory diseases and Alzheimer’s Disease^33^. Among the proteins associated with DNAm PhenoAge, S100A8, S100A9, B2M, and ORM1;ORM2 have previously been shown to be associated with accelerated aging and neuroinflammation^34–36^

These results demonstrate that CXR-Age may identify patients with subclinical functional decline (cardiopulmonary and frailty). A potential clinical use of CXR-Age is to improve medical decisions currently predicated on chronological age^1,37,38^ including clinical risk scores and screening/prevention guidelines. Replacing chronological with CXR-Age may better capture an individual’s current functional status and susceptibility to future insult. Chest radiographs are widely performed in routine clinical care, as a CXR is ordered in 34% of emergency department visits.^39^ CXR-Age could leverage these widely available tests in an “opportunistic screening” approach in which CXRs obtained for other purposes are automatically fed to the CXR-Age model to determine biological age. This biological age can then inform decisions around disease screening and risk assessment that currently rely on chronological age.

The PBHS data are unique in that it is a diverse sample with a great breadth of available data including imaging, epigenetics, and a broad array of functional measures; however, our study has several limitations. We used Horvath Age and DNAm PhenoAge as representative first- and second-generation epigenetic clocks, respectively; however, newer epigenetic clocks are available (e.g., GrimAge^40^) that may have a stronger association with functional outcomes. In addition, several other -omics-based biologic age clocks have been developed, and we aim to study these in future work^6,41^. Longitudinal data (including serial CXRs) were unavailable in this cohort so associations with incident outcomes were not assessed, and it is unclear whether the age acceleration measures predate subclinical cardiopulmonary disease and frailty. While this dataset represents four sites from two states, the results may not generalize to other domestic or international populations. This was a retrospective, observational study, so the causal mechanism linking aging clocks with functional outcomes must still be assessed in prospective studies. Socioeconomic measures and detailed smoking histories were unavailable in this cohort and were not included in multivariable adjustment which may introduce residual confounding. The CXR-Age model was developed in cancer screening trial data in adults 55-74 years; this may explain the attenuated associations in adults younger than 40 years. Lastly, deep learning models are a black box^42^ in that it is unclear what aspects of the image are used to arrive at a prediction. In some cases, we observed that those with the highest age acceleration tended to have less clear lung fields, and we previously found heatmap-based activations^43^ in the mediastinum, aortic knob, and lungs.^20^ however, future interpretability studies are necessary to better understand which image features are important to assess biological age.

In summary, we found that CXR-Age was strongly associated with worse cardiopulmonary function and frailty. Similar associations were found for a second-generation epigenetic clock in older adults but were attenuated in younger age groups. Proteomic analyses revealed novel associations of image and epigenetic aging clocks with proteins linked to inflammation including a shared association with CDH-13 abundance. Future work will assess associations of these and other aging clocks with age-related disease incidence and whether these biological aging measures can improve medical decision-making.

## METHODS

### Study Cohort

Our cohort consisted of 2,097 participants with posterior-anterior CXRs from the Project Baseline Health Study^22^ (sponsored by Verily Life Sciences) (ClinicalTrials.gov Identifier: NCT03154346), a prospective cohort study of individuals from four US sites enriched for cardiovascular and cardiometabolic disease risk factors (Table 1). A subset (N=1,294) of participants had serum methylation data which we used to calculate DNAm PhenoAge and Horvath DNA methylation clocks, respectively,^12,15,20^ and a further subset (N=957) had plasma proteomics measured. This study was approved by the Mass General Brigham institutional review board with a waiver of informed consent for retrospective analysis of deidentified data.

### CXR-Age and chest radiographs

PBHS participants had routine posterior-anterior chest radiographs, and these images were available in standard DICOM format. We preprocessed each CXR using histogram normalization^44^ due to the relatively low contrast in the original CXRs. We manually reviewed all CXRs to ensure correct orientation and quality. No images were removed for poor quality. We then applied the existing CXR-Age model as originally described.^20^ without any modification or fine-tuning to each image to calculate CXR-Age (https://github.com/circ-ml/CXR-Age). Then, we calculated “CXR-Age acceleration” by regressing chronological age and sex on CXR-Age and subtracting this regression from the raw CXR-Age value. For example, a 55-year-old female with a CXR-Age acceleration of 2 years indicates her CXR appears 2 years older than the average CXR for 55-year-old females.

### Measures of cardiopulmonary disease and frailty

We considered measures of cardiovascular disease (Agatston coronary artery calcium [CAC] score from calcium scoring computed tomography (CT)^45^ and 10-year atherosclerotic cardiovascular disease [ASCVD] risk score^46^ calculated from risk factors like cholesterol and blood pressure), pulmonary function (FEV1/FVC ratio, DLCO % predicted, FEV1 % predicted, peak expiratory flow % predicted), physical function/frailty (six-minute walking test, hand-grip strength, sit-stand test, leg balance test, and walking speed), and disability scores (Sheehan and WHODAS 2.0).^47,48^ PBHS participants self-reported risk factors including BMI, smoking frequency, sex, and age to Project Baseline study personnel.

### DNA Methylation Data

PBHS collected Genomic DNA (gDNA) from stored frozen whole blood. Then, they quantified the extracted gDNA using the Quant-iT PicoGreen dsDNA Assay Kit (Invitrogen, Waltham, MA) and ran it on a CLARIOstar microplate reader (BMG LABTECH, Ortenberg, Germany). PBHS processed Bisulfite-converted ssDNA for each sample and control through an automated version of the standard Illumina Infinium MethylationEPIC microarray assay protocol (Illumina Inc., San Diego, CA, USA). PBHS scanned BeadChips on an Illumina iScan (Illumina Inc., San Diego, CA, USA) which generated IDAT files for downstream computational processing. With the resulting CpG site data, we calculated Horvath age and DNAm PhenoAge using previously described methods.^12,15^ We replaced missing CpG site values needed to calculate epigenetic ages with their median values provided in their respective original manuscripts. We calculated epigenetic age acceleration for Horvath age and DNAm PhenoAge using the same method as for CXR-Age acceleration (see CXR-Age and chest radiographs, above)

### Protein Abundance Data

Plasma protein abundance data were available for 957 participants in the Project Baseline Health Study. We used these data to hypothesize potential biological mechanisms of CXR-Age and epigenetic age acceleration. PBHS prepared each plasma protein sample by first doing microflow high-resolution liquid chromatography-mass spectrometry. Then, they converted this raw data to protein abundances through the use of Dia-NN v1.8.1 (https://github.com/vdemichev/DiaNN).^49^ They implemented various quality control steps. At the end, PBHS detected 289 proteins across all patient plasma samples. Microbial proteins, contaminants and Ig variable chain proteins are not included in the analysis. PBHS collected protein abundance data in arbitrary units (AU).

### Statistical Analysis

We assessed the association between CXR-Age and DNA Methylation-age with continuous outcomes using linear regression and reported the results per 1-year increase in age acceleration. We applied a log transformation to CAC score and ASCVD risk score to achieve normality. We adjusted all analyses for chronological age, body mass index (BMI), sex, smoking status, race, and recruiting site. We used a p-value threshold of 0.05 for cardiopulmonary and frailty outcomes. We used a Bonferroni corrected p-value threshold of 1.7e-4 for proteomic analysis to account for the 289 proteins analyzed.

## Data Availability

All data used in the present study may be available with prior approval from Project Baseline Health Study

## ACKNOWLEDGMENTS

The authors wish to thank Project Baseline Health Study participants and study sites.

## Data Sharing Statement

The deidentified PBHS data corresponding to this study are available upon request for the purpose of examining its reproducibility. Requests are subject to approval by PBHS governance.

## Ethics Statement

The study was approved by the Duke University and Stanford University Institutional Review Boards. Informed consent was obtained from all participants enrolled in the Project Baseline Health Study in accordance with the Declaration of Helsinki.

## REFERENCES

1. Arnett DK, Blumenthal RS, Albert MA, et al. 2019 ACC/AHA Guideline on the Primary Prevention of Cardiovascular Disease: A Report of the American College of Cardiology/American Heart Association Task Force on Clinical Practice Guidelines. Circulation. 2019;140(11):e596-e646. doi:10.1161/CIR.0000000000000678

2. Chin J, Baldwin J, Evans M, Long K, Li C, Mukherjee D. Screening for Lung Cancer with Low Dose Computed Tomography. Published online February 10, 2022. https://www.cms.gov/medicare-coverage-database/view/ncacal-decision-memo.aspx?proposed=N&ncaid=304

3. Elliott ML, Caspi A, Houts RM, et al. Disparities in the pace of biological aging among midlife adults of the same chronological age have implications for future frailty risk and policy. Nat Aging. 2021;1(3):295–308. doi:10.1038/s43587-021-00044-4

4. Moqri M, Herzog C, Poganik JR, et al. Biomarkers of Aging for the Identification and Evaluation of Longevity Interventions. Cell. 2023;186(18):3758–3775. doi:10.1016/j.cell.2023.08.003

5. Duan R, Fu Q, Sun Y, Li Q. Epigenetic clock: A promising biomarker and practical tool in aging. Ageing Res Rev. 2022;81:101743. doi:10.1016/j.arr.2022.101743

6. Argentieri MA, Xiao S, Bennett D, et al. Proteomic aging clock predicts mortality and risk of common age-related diseases in diverse populations. Nat Med. Published online August 8, 2024:1–11. doi:10.1038/s41591-024-03164-7

7. Sanders JL, Newman AB. Telomere length in epidemiology: a biomarker of aging, age-related disease, both, or neither? Epidemiol Rev. 2013;35(1):112–131. doi:10.1093/epirev/mxs008

8. Bohannon RW. Grip Strength: An Indispensable Biomarker For Older Adults. Clin Interv Aging. 2019;14:1681–1691. doi:10.2147/CIA.S194543

9. Je H, Da C, Pa R, Dh P. Age-related changes in speed of walking. Med Sci Sports Exerc. 1988;20(2). doi:10.1249/00005768-198820020-00010

10. de Hollander EL, Bemelmans WJ, Boshuizen HC, et al. The association between waist circumference and risk of mortality considering body mass index in 65- to 74-year-olds: a meta-analysis of 29 cohorts involving more than 58 000 elderly persons. Int J Epidemiol. 2012;41(3):805–817. doi:10.1093/ije/dys008

11. Ponti F, Santoro A, Mercatelli D, et al. Aging and Imaging Assessment of Body Composition: From Fat to Facts. Front Endocrinol. 2020;10:861. doi:10.3389/fendo.2019.00861

12. Horvath S. DNA methylation age of human tissues and cell types. Genome Biol. 2013;14(10):R115. doi:10.1186/gb-2013-14-10-r115

13. Hannum G, Guinney J, Zhao L, et al. Genome-wide methylation profiles reveal quantitative views of human aging rates. Mol Cell. 2013;49(2):359–367. doi:10.1016/j.molcel.2012.10.016

14. Fransquet PD, Wrigglesworth J, Woods RL, Ernst ME, Ryan J. The epigenetic clock as a predictor of disease and mortality risk: a systematic review and meta-analysis. Clin Epigenetics. 2019;11(1):62. doi:10.1186/s13148-019-0656-7

15. Levine ME, Lu AT, Quach A, et al. An epigenetic biomarker of aging for lifespan and healthspan. Aging. 2018;10(4):573–591. doi:10.18632/aging.101414

16. Gialluisi A, Santoro A, Tirozzi A, et al. Epidemiological and genetic overlap among biological aging clocks: New challenges in biogerontology. Ageing Res Rev. 2021;72:101502. doi:10.1016/j.arr.2021.101502

17. Porter HL, Brown CA, Roopnarinesingh X, et al. Many chronological aging clocks can be found throughout the epigenome: Implications for quantifying biological aging. Aging Cell. 2021;20(11):e13492. doi:10.1111/acel.13492

18. Bell CG, Lowe R, Adams PD, et al. DNA methylation aging clocks: challenges and recommendations. Genome Biol. 2019;20(1):249. doi:10.1186/s13059-019-1824-y

19. Smith-Bindman R, Kwan ML, Marlow EC, et al. Trends in Use of Medical Imaging in US Health Care Systems and in Ontario, Canada, 2000-2016. JAMA. 2019;322(9):843–856. doi:10.1001/jama.2019.11456

20. Raghu VK, Weiss J, Hoffmann U, Aerts HJWL, Lu MT. Deep Learning to Estimate Biological Age From Chest Radiographs. JACC Cardiovasc Imaging. 2021;14(11):2226–2236. doi:10.1016/j.jcmg.2021.01.008

21. Lee JH, Lee D, Lu MT, et al. External Testing of a Deep Learning Model to Estimate Biologic Age Using Chest Radiographs. Radiol Artif Intell. 2024;6(5):e230433. doi:10.1148/ryai.230433

22. Arges K, Assimes T, Bajaj V, et al. The Project Baseline Health Study: a step towards a broader mission to map human health. Npj Digit Med. 2020;3(1):1–10. doi:10.1038/s41746-020-0290-y

23. Goff DC, Lloyd-Jones DM, Bennett G, et al. 2013 ACC/AHA guideline on the assessment of cardiovascular risk: a report of the American College of Cardiology/American Heart Association Task Force on Practice Guidelines. Circulation. 2014;129(25 Suppl 2):S49–73. doi:10.1161/01.cir.0000437741.48606.98

24. Joyce BT, Gao T, Zheng Y, et al. Epigenetic Age Acceleration Reflects Long-Term Cardiovascular Health. Circ Res. 2021;129(8):770–781. doi:10.1161/CIRCRESAHA.121.318965

25. Sánchez-Cabo F, Fuster V, Silla-Castro JC, et al. Subclinical atherosclerosis and accelerated epigenetic age mediated by inflammation: a multi-omics study. Eur Heart J. 2023;44(29):2698–2709. doi:10.1093/eurheartj/ehad361

26. Phyo AZZ, Espinoza SE, Murray AM, et al. Epigenetic age acceleration and the risk of frailty, and persistent activities of daily living (ADL) disability. Age Ageing. 2024;53(6):afae127. doi:10.1093/ageing/afae127

27. McCrory C, Fiorito G, Hernandez B, et al. GrimAge Outperforms Other Epigenetic Clocks in the Prediction of Age-Related Clinical Phenotypes and All-Cause Mortality. J Gerontol Ser A. 2021;76(5):741–749. doi:10.1093/gerona/glaa286

28. The relationship between the epigenetic aging biomarker “grimage” and lung function in both the airway and blood of people living with HIV: An observational cohort study - eBioMedicine. Accessed December 16, 2024. https://www.thelancet.com/journals/ebiom/article/PIIS2352-3964(22)00388-7/fulltext

29. Breen M, Nwanaji-Enwerem JC, Karrasch S, et al. Accelerated epigenetic aging as a risk factor for chronic obstructive pulmonary disease and decreased lung function in two prospective cohort studies. Aging. 2020;12(16):16539–16554. doi:10.18632/aging.103784

30. Hillary RF, Stevenson AJ, McCartney DL, et al. Epigenetic measures of ageing predict the prevalence and incidence of leading causes of death and disease burden. Clin Epigenetics. 2020;12:115. doi:10.1186/s13148-020-00905-6

31. Rivero O, Selten MM, Sich S, et al. Cadherin-13, a risk gene for ADHD and comorbid disorders, impacts GABAergic function in hippocampus and cognition. Transl Psychiatry. 2015;5(10):e655. doi:10.1038/tp.2015.152

32. Rassart E, Desmarais F, Najyb O, Bergeron KF, Mounier C. Apolipoprotein D. Gene. 2020;756:144874. doi:10.1016/j.gene.2020.144874

33. SAA2 serum amyloid A2 [Homo sapiens (human)] - Gene - NCBI. Accessed December 16, 2024. https://www.ncbi.nlm.nih.gov/gene?Db=gene&Cmd=DetailsSearch&Term=6289

34. Wang S, Song R, Wang Z, Jing Z, Wang S, Ma J. S100A8/A9 in Inflammation. Front Immunol. 2018;9. doi:10.3389/fimmu.2018.01298

35. Zhong Q, Zou Y, Liu H, et al. Toll-like receptor 4 deficiency ameliorates β2-microglobulin induced age-related cognition decline due to neuroinflammation in mice. Mol Brain. 2020;13(1):20. doi:10.1186/s13041-020-0559-8

36. Ligresti G, Aplin AC, Dunn BE, Morishita A, Nicosia RF. The Acute Phase Reactant Orosomucoid-1 Is a Bimodal Regulator of Angiogenesis with Time- and Context-Dependent Inhibitory and Stimulatory Properties. PLOS ONE. 2012;7(8):e41387. doi:10.1371/journal.pone.0041387

37. US Preventive Services Task Force, Krist AH, Davidson KW, et al. Screening for Lung Cancer: US Preventive Services Task Force Recommendation Statement. JAMA. 2021;325(10):962. doi:10.1001/jama.2021.1117

38. Validation of risk stratification schemes for predicting stroke and thromboembolism in patients with atrial fibrillation: nationwide cohort study | The BMJ. Accessed December 16, 2024. https://www.bmj.com/content/342/bmj.d124

39. Fatihoglu E, Aydin S, Gokharman FD, Ece B, Kosar PN. X-ray Use in Chest Imaging in Emergency Department on the Basis of Cost and Effectiveness. Acad Radiol. 2016;23(10):1239–1245. doi:10.1016/j.acra.2016.05.008

40. DNA methylation GrimAge strongly predicts lifespan and healthspan - PMC. Accessed December 16, 2024. https://pmc.ncbi.nlm.nih.gov/articles/PMC6366976/

41. Krištić J, Vučković F, Menni C, et al. Glycans are a novel biomarker of chronological and biological ages. J Gerontol A Biol Sci Med Sci. 2014;69(7):779–789. doi:10.1093/gerona/glt190

42. Adebayo J, Gilmer J, Muelly M, Goodfellow I, Hardt M, Kim B. Sanity Checks for Saliency Maps. Published online November 6, 2020. doi:10.48550/arXiv.1810.03292

43. Selvaraju RR, Cogswell M, Das A, Vedantam R, Parikh D, Batra D. Grad-CAM: Visual Explanations from Deep Networks via Gradient-based Localization. Int J Comput Vis. 2020;128(2):336–359. doi:10.1007/s11263-019-01228-7

44. Giełczyk A, Marciniak A, Tarczewska M, Lutowski Z. Pre-processing methods in chest X-ray image classification. PLoS ONE. 2022;17(4):e0265949. doi:10.1371/journal.pone.0265949

45. Haddad F, Cauwenberghs N, Daubert MA, et al. Association of left ventricular diastolic function with coronary artery calcium score: A Project Baseline Health Study. J Cardiovasc Comput Tomogr. 2022;16(6):498–508. doi:10.1016/j.jcct.2022.06.003

46. Stone NJ, Robinson JG, Lichtenstein AH, et al. 2013 ACC/AHA Guideline on the Treatment of Blood Cholesterol to Reduce Atherosclerotic Cardiovascular Risk in Adults: A Report of the American College of Cardiology/American Heart Association Task Force on Practice Guidelines. J Am Coll Cardiol. 2014;63(25, Part B):2889–2934. doi:10.1016/j.jacc.2013.11.002

47. Sheehan DV, Harnett-Sheehan K, Raj BA. The measurement of disability. Int Clin Psychopharmacol. 1996;11 Suppl 3:89–95. doi:10.1097/00004850-199606003-00015

48. WHO Disability Assessment Schedule (WHODAS 2.0). Accessed December 16, 2024. https://www.who.int/standards/classifications/international-classification-of-functioning-disability-and-health/who-disability-assessment-schedule

49. Demichev V, Messner CB, Vernardis SI, Lilley KS, Ralser M. DIA-NN: neural networks and interference correction enable deep proteome coverage in high throughput. Nat Methods. 2020;17(1):41–44. doi:10.1038/s41592-019-0638-x

