## Supplemental Material for "Deep Learning Chest X-Ray Age, Epigenetic Aging Clocks and Associations with Age-Related Subclinical Disease in the Project Baseline Health Study"

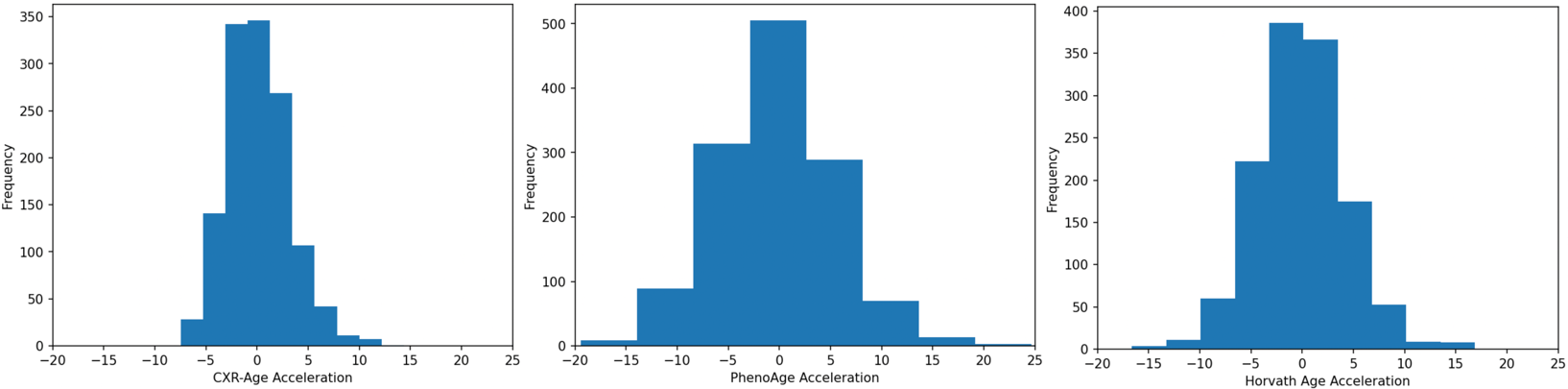

**Supplemental Figure 1. Distribution of CXR-Age Acceleration and Epigenetic Age Acceleration**

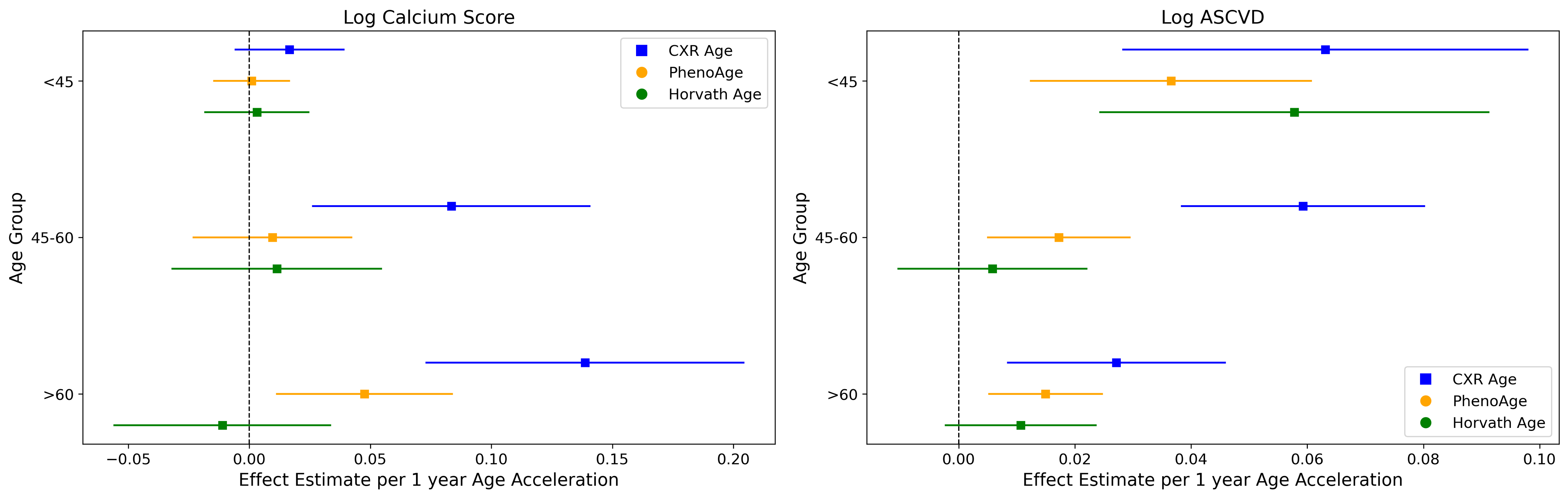

**Supplemental Figure 2. Association of Cardiac Outcomes with CXR-Age and Epigenetic Aging Clocks Stratified by Chronological Age (<45 years, 45-60, >60).** Outcomes include log calcium score and the log ASCVD risk score. Effect estimates and 95% CI per 1-year for each age acceleration metric are provided on the x-axis.

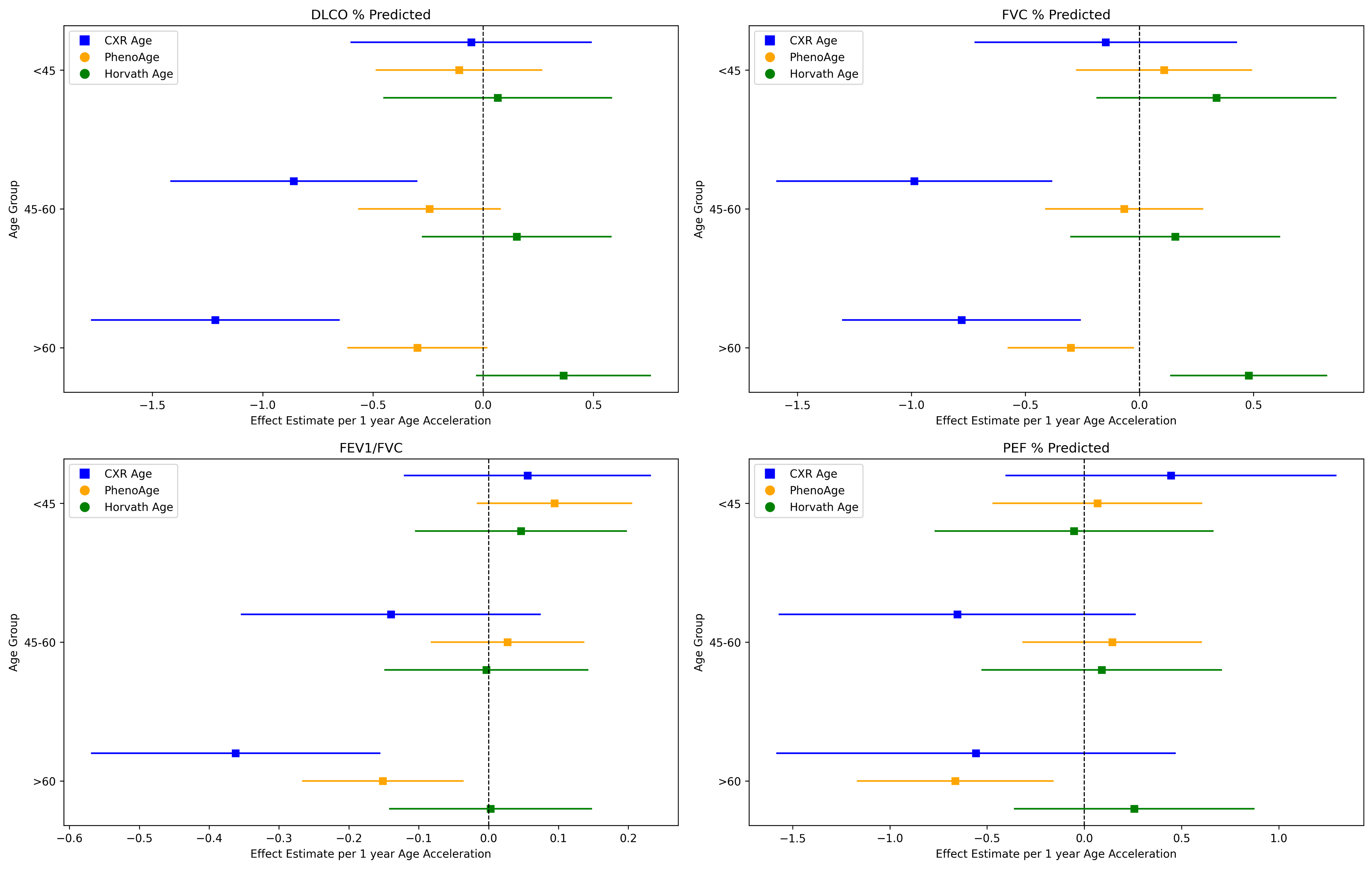

**Supplemental Figure 3. Association of Lung Function Tests with CXR-Age and Epigenetic Aging Clocks Stratified by Chronological Age (<45 years,45-60, >60).** The lung function tests include DLCO% predicted, FVC% predicted, FEV1/FVC ratio, and PEF % predicted. Effect estimates and 95% CI per 1-year for each age acceleration metric are provided on the x-axis.

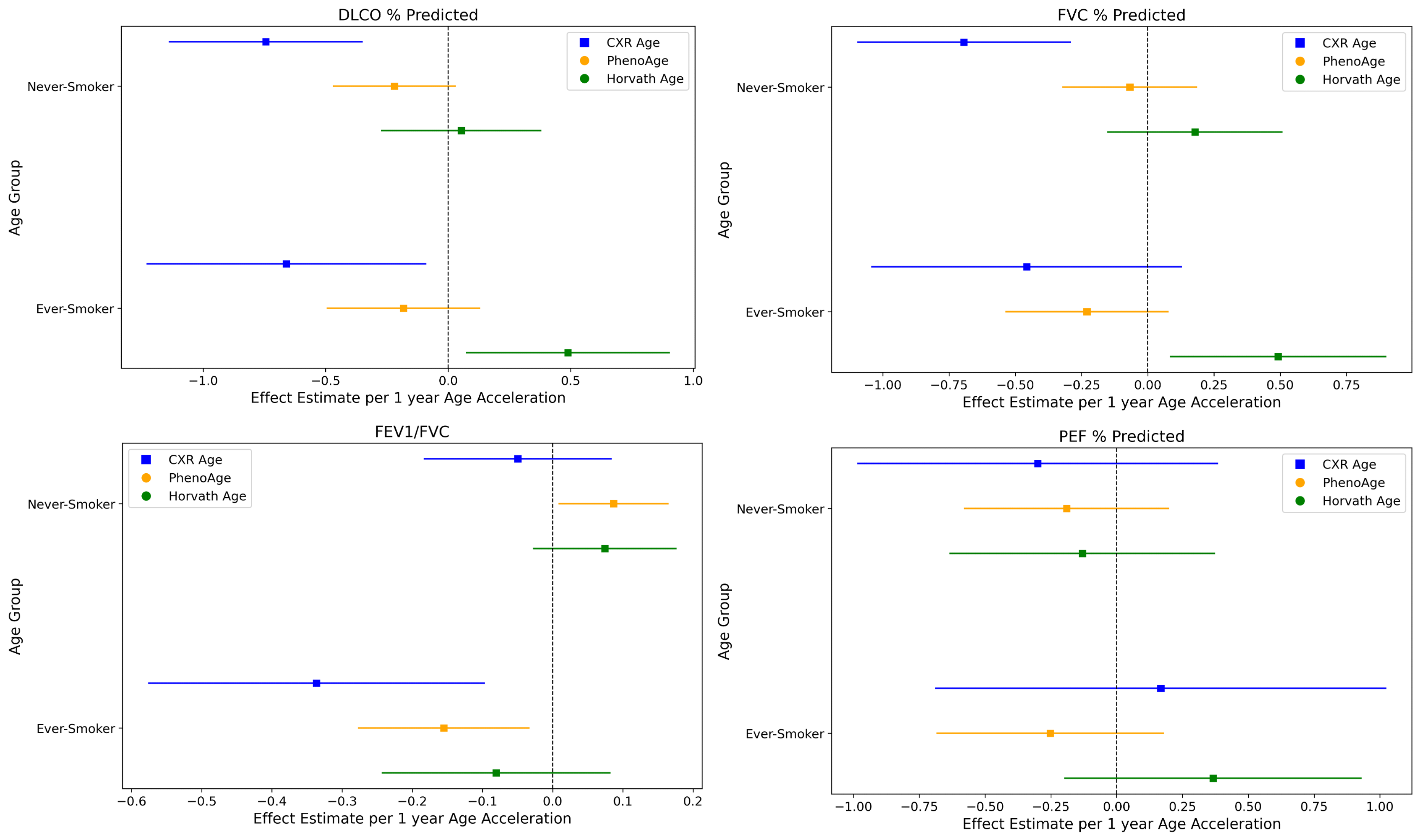

**Supplemental Figure 4. Association of Lung Function Tests with CXR-Age and Epigenetic Aging Clocks Stratified by Smoking Status.** The lung function tests include DLCO% predicted, FVC% predicted, FEV1/FVC ratio, and PEF % predicted. Effect estimates and 95% CI per 1-year for each age acceleration metric are provided on the x-axis.

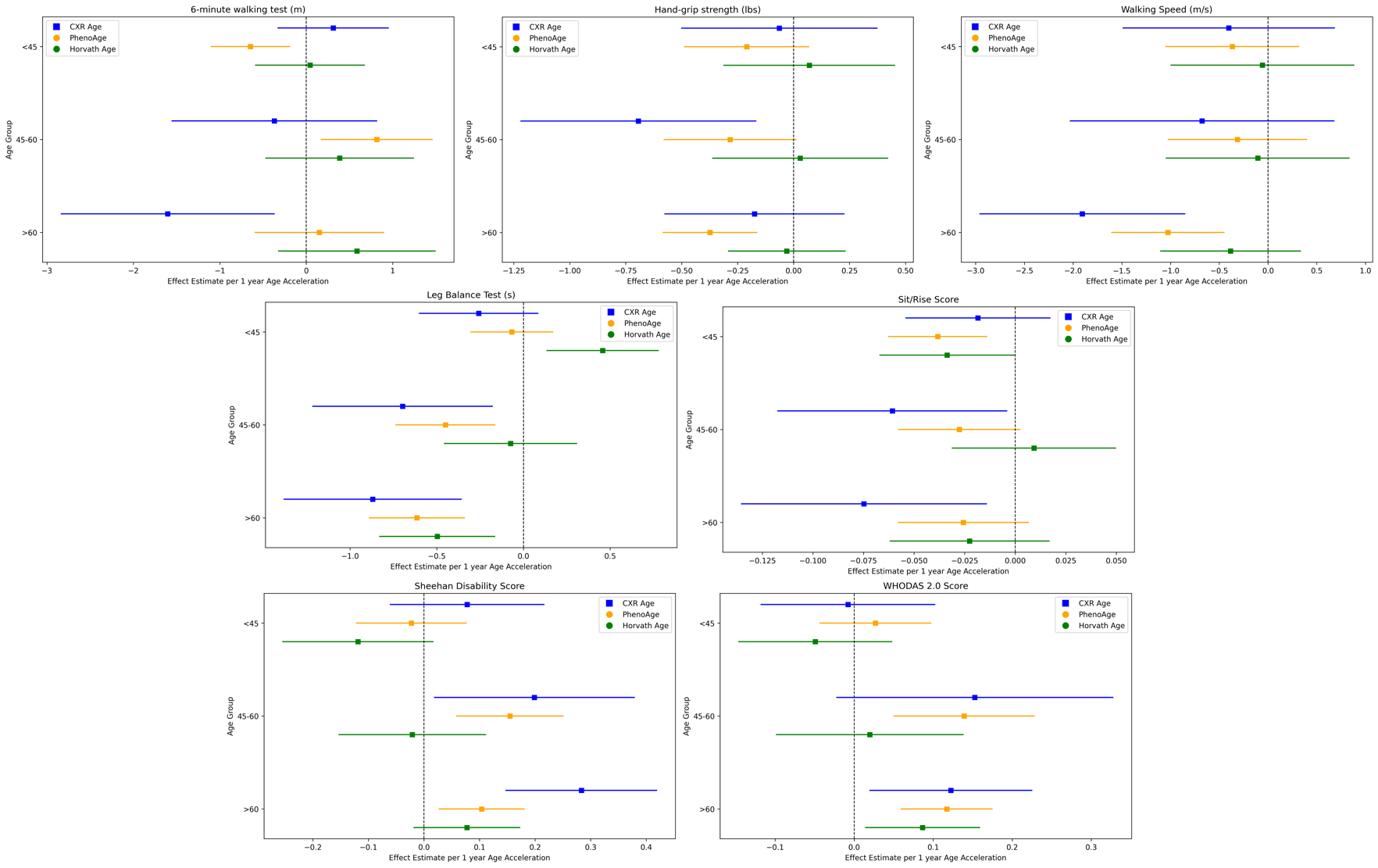
**Supplemental Figure 5. Association of Frailty Outcomes and with CXR-Age and Epigenetic Aging Clocks Stratified by Chronological Age (<45 years, 45-60, >60).** Outcomes include the 6-minute walking test (m), hand-grip strength (lbs), walking speed (m/s), leg balance test (s), sit/rise score, Sheehan disability score, and WHODAS 2.0 score. Effect estimates and 95% CI per 1-year for each age acceleration metric are provided on the x-axis.

| **Outcome** | **CXR-Age Acceleration** | **PhenoAge Acceleration** | **Horvath Age Acceleration** |
| --- | --- | --- | --- |
| Log CACS Adjusted | 0.09[0.06 - 0.13] | 0.03[0.1 - 0.05] | -0.01[-0.04 - 0.02] |
| Log CACS Unadjusted | 0.11[0.07 - 0.14] | 0.03[0.01 - 0.06] | -0.0[-0.04 - 0.03] |
| Log ASCVD Adjusted | 0.04[0.01 - 0.07] | 0.0[-0.01 - 0.01] | 0.0[-0.01 - 0.01] |
| Log ASCVD Unadjusted | 0.06[0.03 - 0.09] | 0.02[0.0 - 0.04] | 0.01[-0.02 - 0.03] |
| FEV1/FVC Adjusted | -0.17[-0.29 - -0.05] | -0.02[-0.08 - 0.05] | 0.01[-0.08 - 0.09] |
| FEV1/FVC Unadjusted | -0.01[-0.13 - 0.11] | 0.01[-0.06 - 0.08] | 0.02[-0.07 - 0.12] |
| DLCO % Predicted Adjusted | -0.74[-1.06 - -0.42] | -0.23[-0.42 - -0.03] | 0.22[-0.03 - 0.48] |
| DLCO % Predicted Unadjusted | -1.3[-1.64 - -0.96] | -0.45[-0.65 - -0.25] | 0.21[-0.07 - 0.49] |
| FVC % Predicted Adjusted | -0.63[-0.96 - -0.3] | -0.15[-0.35 - 0.04] | 0.28[0.02 - 0.53] |
| FVC % Predicted Unadjusted | -0.98[-1.29 - -0.67] | -0.29[-0.49 - -0.1] | 0.23[-0.03 - 0.49] |
| PEF % Predicted Adjusted | -0.1[-0.63 - 0.44] | -0.21[-0.5 - 0.07] | 0.06[-0.31 - 0.44] |
| PEF % Predicted Unadjusted | -0.26[-0.78 - 0.25] | -0.18[-0.47 - 0.11] | 0.14[-0.25 - 0.53] |
| 6-minute walking test (m) Adjusted | -0.61[-1.19 - -0.03] | 0.02[-0.34 - 0.38] | 0.45[-0.02 - 0.92] |
| 6-minute walking test (m) Unadjusted | -3.1[-4.3 - -1.89] | -0.6[-1.35 - 0.15] | 0.13[-0.88 - 1.14] |
| Hand-grip strength (lbs) Adjusted | -0.3[-0.56 - -0.04] | -0.35[-0.5 - -0.21] | -0.01[-0.2 - 0.19] |
| Hand-grip strength (lbs) Unadjusted | -0.12[-0.48 - 0.23] | -0.25[-0.46 - -0.03] | 0.04[-0.25 - 0.33] |
| Sit/Rise Score Adjusted | -0.05[-0.08 - -0.02] | -0.03[-0.05 - -0.01] | -0.01[-0.03 - 0.01] |
| Sit/Rise Score Unadjusted | -0.12[-0.16 - -0.09] | -0.06[-0.08 - -0.04] | -0.03[-0.06 - -0.0] |
| Leg Balance Test (s) Adjusted | -0.61[-0.87 - -0.35] | -0.45[-0.6 - -0.3] | -0.1[-0.3 - 0.09] |
| Leg Balance Test (s) Unadjusted | -1.43[-1.75 - -1.11] | -0.76[-0.96 - -0.57] | -0.25[-0.52 - 0.01] |
| Walking Speed (cm/s) Adjusted | -1.1[-1.77 - -0.44] | -0.64[-1.01 - -0.27] | -0.2[-0.69 - 0.28] |
| Walking Speed (cm/s) Unadjusted | -2.36[-3.02 - -1.7] | -1.22[-1.61 - -0.83] | -0.47[-1.0 - 0.06] |
| WHODAS 2.0 Score Adjusted | 0.08[0.01 - 0.16] | 0.1[0.06 - 0.14] | 0.03[-0.02 - 0.09] |
| WHODAS 2.0 Score Unadjusted | 0.23[0.16 - 0.3] | 0.16[0.12 - 0.2] | 0.06[-0.0 - 0.11] |
| Sheehan Disability Score Adjusted | 0.16[0.07- 0.25] | 0.06[0.0 - 0.11] | -0.04[-0.11 - 0.03] |
| Sheehan Disability Score Unadjusted | 0.17[0.08 - 0.26] | 0.11[0.05 - 0.16] | -0.02[-0.09 - 0.05] |

**Supplemental Table 1. Comparison of unadjusted and adjusted associations between cardiovascular, lung function, and frailty outcomes and biological ages (CXR-Age, PhenoAge, Horvath Age).** Effect estimates for a 1-year increase in age acceleration. Adjusted results control for chronological age, sex, race, BMI, smoking status, and recruitment site.
